# Cross-sectional Study on Sex Disparities among Japanese and Korean University School of Pharmacy Professors

**DOI:** 10.1101/2025.01.14.25320528

**Authors:** Hyunseo Soh, Eri Ohno, Mayu Hashimoto, Euna Han, Hideki Maeda

## Abstract

Japan and South Korea are ranked low in the Gender Gap Index, and the ratio of female faculty members at universities is low. The ratio of female professors at universities is assumed to be lower than the ratio of female medical professionals; however, the details remain unknown. Consequently, this study aimed to examine sex disparity among professors in the pharmacy departments of universities in Japan and South Korea. Furthermore, the characteristics of Japanese and South Korean pharmacy departments were examined to explore the differences and sex disparity.

This study was conducted based on publicly available information from the websites of each university’s pharmacy department as of November 1, 2023. Sex was determined from first names or photographs. This study was conducted following the STROBE guidelines, and no ethical review or consent was required.

This study included 73 universities in Japan and 35 universities in South Korea. The proportion of female professors in university pharmacy departments was 12.5% in Japan and 32.9% in South Korea. A total of 1,965 pharmacy professors were identified in Japan and South Korea. Of these, 1,527 were from Japan and 438 were from South Korea. The estimated ratio of professors to million individuals in Japan was 12.28, compared to 8.51 in South Korea. This indicated that Japan had approximately 1.4 times as many professors as South Korea after correction for population.

The proportion of female professors in both countries was <50%. Japan had significantly fewer female professors than South Korea, suggesting that women’s advancement in university pharmacy departments was lagging in Japan. Additionally, South Korea’s higher number of female professors specializing in clinical research may suggest that women with clinical practice experience in hospitals or pharmacies were becoming university professors.

## 1. Introduction

Sex disparities have been a significant issue in various professional fields [1], particularly in Japan and South Korea. Both countries have a historical background where Confucianism shaped perspectives on life, lifestyle, and ethics, which contributed to their positions as lower-ranked countries in the Gender Gap Index [2,3]. The low proportion of female faculty members in universities has been a long-standing issue, and although the number of female educators is gradually increasing, a sex gap remains [4]. Recent survey results indicated that the proportion of female faculty members decreases as academic rank increases [5]. As developed nations, Japan and South Korea are qualified to promote the advancement of science and are actively enhancing women’s career progression in scientific fields. However, it is presumed that a dearth of capable female researchers or educators in authoritative positions impedes women’s career advancement. Furthermore, female researchers tend to eschew competitive situations, frequently undervalue their capabilities, and may be disinclined to pursue leadership roles [6]. To expand women’s participation and promote diversity in all areas of society, the Japanese government initiated the forward-thinking program, the Empowerment Network, in July 2018 [7]. This initiative set a goal of achieving at least 30% female representation in leadership roles across all sectors of society, including politics, national and local civil service, the private sector, education, and research, by 2020. Similar efforts have been undertaken in South Korea [8].

A major aim of university pharmacy departments is to train pharmacists. However, a reverse sex gap is evident in this field, with a high proportion of women among pharmacists, and more female than male students in university pharmacy departments [9,10]. Despite this, few studies have examined sex disparity among university pharmacy faculties, and several aspects remain unclear. A previous study on sex disparities among pharmacy school professors in Japan and Korea has been conducted and the preliminary results have been reported [11]. However, the detailed results have not yet been presented.

This study further examined sex disparity among professors in university pharmacy departments in Japan and South Korea as of November 2023. It examined whether sex disparity exists among pharmacy professors in both countries, as well as the details of this disparity. Additionally, this study explored each country’s university pharmacy departments’ characteristics to determine the differences concerning the backgrounds of university pharmacy departments in Japan and South Korea.

## 2. Materials and Methods

### 2.1. Study design

This study employed a cross-sectional design.

### 2.2. Data abstraction

This study was conducted following the Strengthening the Reporting of Observational Studies in Epidemiology (STROBE) reporting guidelines [12] for cross-sectional studies. Data collection was independently conducted by two of the authors. In cases where discrepancies or conflicts occurred, consultations were held with the third author to reach a consensus. The data were obtained from publicly available information from the websites of each university. This study did not require institutional review board approval or patient informed consent, given that it was based on publicly available information and involved no patient records.

### 2.3. Data collection

#### 2.3.1. Subjects

This study was conducted based on information as of November 1, 2023 (predominantly from university websites). This study focused on professors in the pharmacy departments of universities in Japan and South Korea, excluding university presidents, emeritus professors, and newly established universities with no graduates as of 2023.

#### 2.3.2. Specialty field of each professor

The academic specialties of each professor were determined based on the university and laboratory websites, using the names of the laboratories and their research content. The professors were first categorized into basic and clinical research fields. The subsequent classifications were as follows: 1) Basic Research: Physics, Chemistry, Biology, Hygiene, Pharmacology, and Pharmaceutical Sciences; 2) Clinical Research: Pathology and Drug Therapy, Laws, Systems, and Ethics, and Clinical Training. The criteria for distinguishing between basic and clinical research were based on Japan’s national examination subjects [13].

#### 2.3.3. Background of each university

This study predominantly used information from each university’s website to gather the following details: year of establishment, founder (whether national or private), faculty composition (whether university; offering multiple faculties, or college; offering a single faculty), sex of students (coeducational or women’s university), annual number of student per grade, number of professors (total number of pharmacy faculty professors), 2022 national examination pass rate (for Japan only), and difficulty of admission (for Japan, based on the Kawaijuku standard score for 2022 (https://www.keinet.ne.jp/university/ranking/); for South Korea, based on one of the authors’ university entrance experience).

#### 2.3.4. Determination of sex

The subjects’ sex was determined based on first names or photographs. For example, names that ended with “ko” or “mi,” such as Reiko or Mayumi, were female. In Korea, it can be relatively difficult to distinguish between male and female names; consequently, photographs were predominantly used to make the decision. When these were insufficient for determining sex, additional information such as resumes, curriculum vitae, or personal information available on the universities’ websites was used to ascertain sex. If sex could still not be determined, further steps were taken, such as direct inquiries by phone to the respective university. Regarding judgments based on first names, Japanese names were assessed by native Japanese speakers, and Korean names were assessed by native Korean speakers. Three of the authors are native Japanese speakers, and two are native Korean speakers.

#### 2.3.5. Location of university (city area/regional area)

Regarding Japan, the city area was defined as the designated cities and Tokyo, with all other areas classified as regional.

Regarding South Korea, the city area was defined as Seoul, Gyeonggi-do, and Incheon, with all other areas classified as regional.

#### 2.3.6. Number of students per grade

The number of students per grade referred to the number of departments that offered a pharmacy license.

### 2.4. Statistical analysis

Descriptive statistics were used to examine the universities’ characteristics and pharmacy professors’ sex in Japan and South Korea. A Chi-squared test was used to evaluate the characteristics and sex disparities. All analyses were conducted using R software (Version 4.2.1). The statistical significance level was set at a two-sided *p*=0.05.

## 3. Results

### 3.1. Comparison of university pharmacy departments in Japan and South Korea

This study included 73 Japanese and 35 South Korean universities. No significant differences were observed between Japan and South Korea concerning the backgrounds of the examined universities (national vs. private university), the composition of the faculty (university vs. college), or the location (city area vs. regional area). However, significant differences were observed concerning the establishment year, the annual number of students per grade, and the number of professors. Korean universities were significantly newer, while Japanese universities had significantly larger student enrollment capacities and more professors. Additionally, regarding faculty composition, South Korea had no college pharmacy departments (Table 1).

**Table 1.**
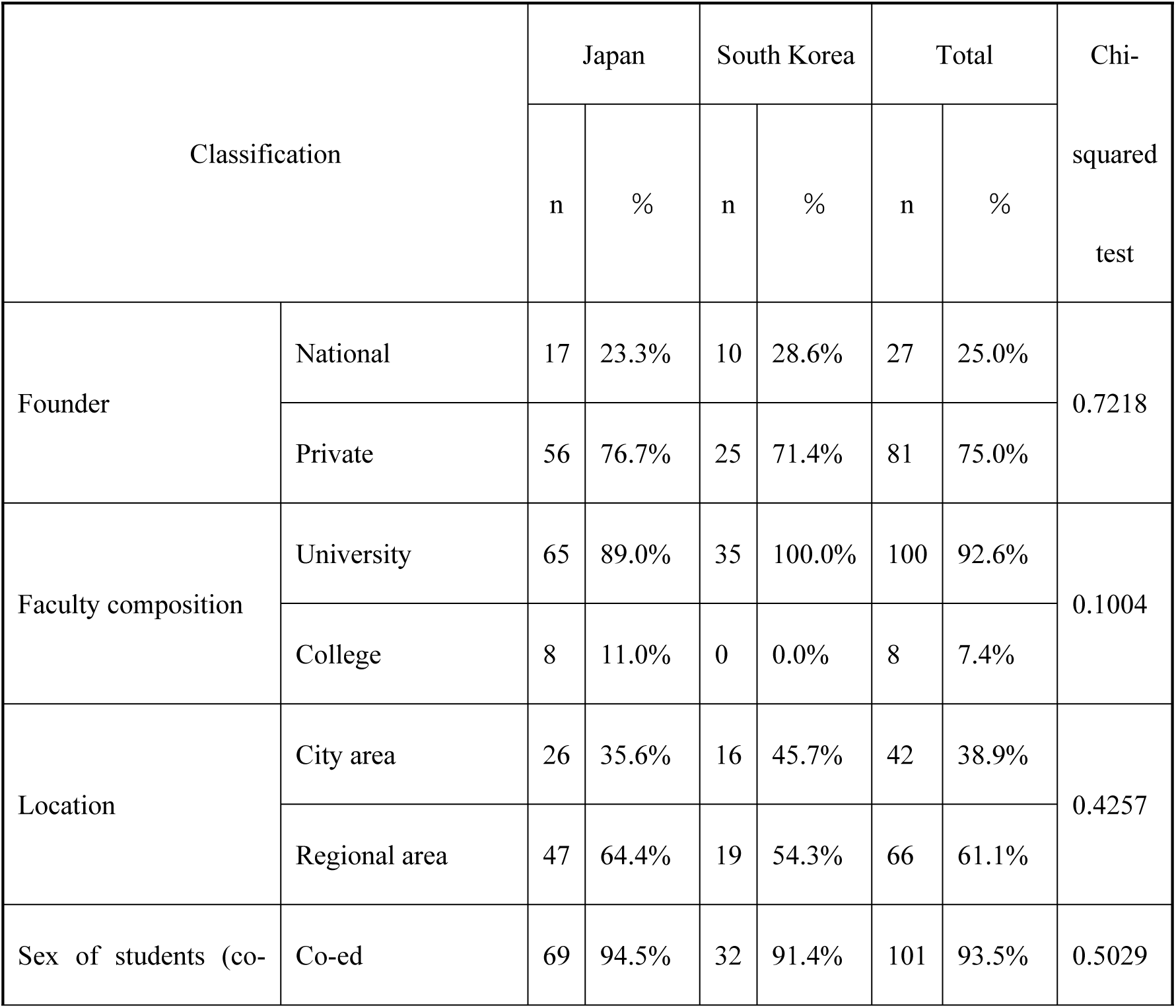
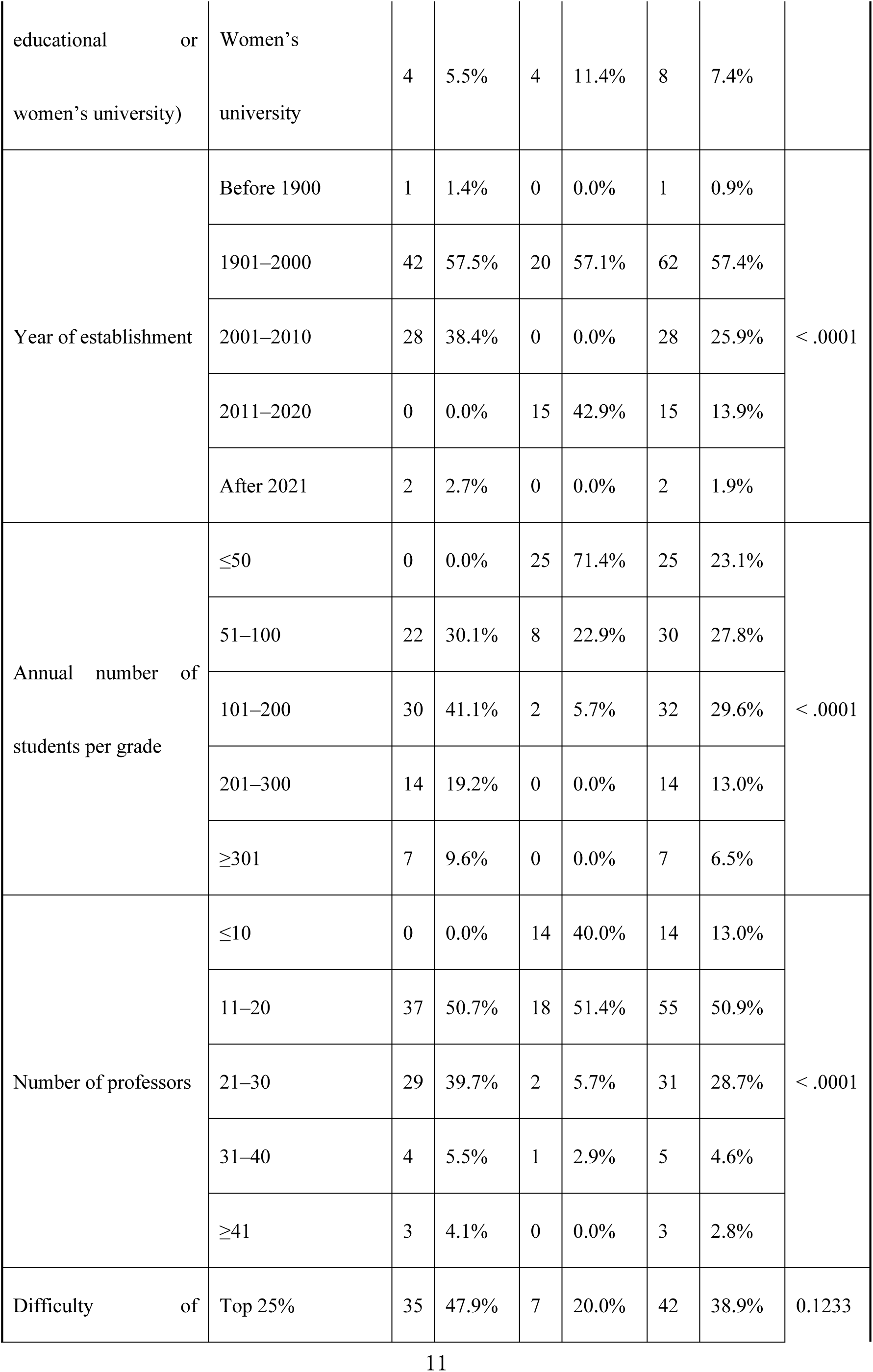
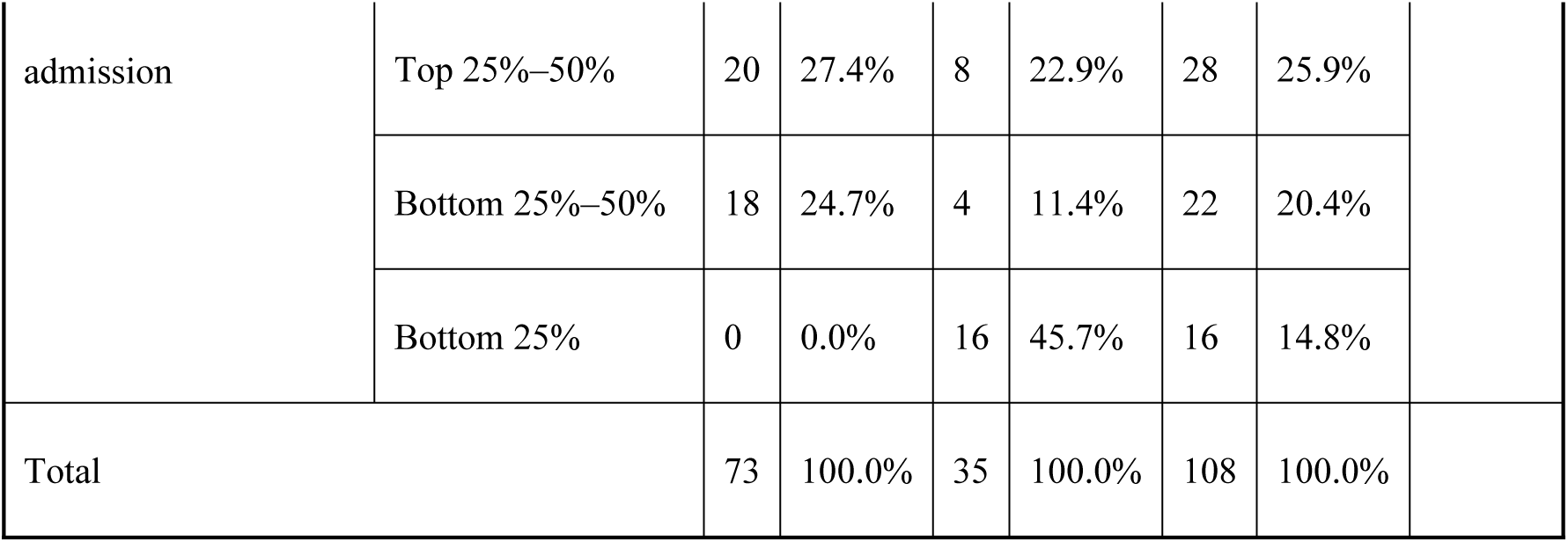
Backgrounds of examined university pharmacy departments

| Classification |  | Japan |  | South Korea |  | Total |  | Chi-squared test |
| --- | --- | --- | --- | --- | --- | --- | --- | --- |
|  |  | n | % | n | % | n | % |  |
| Founder | National | 17 | 23.3% | 10 | 28.6% | 27 | 25.0% | 0.7218 |
|  | Private | 56 | 76.7% | 25 | 71.4% | 81 | 75.0% |  |
| Faculty composition | University | 65 | 89.0% | 35 | 100.0% | 100 | 92.6% | 0.1004 |
|  | College | 8 | 11.0% | 0 | 0.0% | 8 | 7.4% |  |
| Location | City area | 26 | 35.6% | 16 | 45.7% | 42 | 38.9% | 0.4257 |
|  | Regional area | 47 | 64.4% | 19 | 54.3% | 66 | 61.1% |  |
| Sex of students (co- | Co-ed | 69 | 94.5% | 32 | 91.4% | 101 | 93.5% | 0.5029 |
| educational or<br>women's university) | Women's<br>university | 4 | 5.5% | 4 | 11.4% | 8 | 7.4% |  |
| Year of establishment | Before 1900 | 1 | 1.4% | 0 | 0.0% | 1 | 0.9% | < .0001 |
|  | 1901–2000 | 42 | 57.5% | 20 | 57.1% | 62 | 57.4% |  |
|  | 2001–2010 | 28 | 38.4% | 0 | 0.0% | 28 | 25.9% |  |
|  | 2011–2020 | 0 | 0.0% | 15 | 42.9% | 15 | 13.9% |  |
|  | After 2021 | 2 | 2.7% | 0 | 0.0% | 2 | 1.9% |  |
| Annual number of<br>students per grade | ≤50 | 0 | 0.0% | 25 | 71.4% | 25 | 23.1% | < .0001 |
|  | 51–100 | 22 | 30.1% | 8 | 22.9% | 30 | 27.8% |  |
|  | 101–200 | 30 | 41.1% | 2 | 5.7% | 32 | 29.6% |  |
|  | 201–300 | 14 | 19.2% | 0 | 0.0% | 14 | 13.0% |  |
|  | ≥301 | 7 | 9.6% | 0 | 0.0% | 7 | 6.5% |  |
| Number of professors | ≤10 | 0 | 0.0% | 14 | 40.0% | 14 | 13.0% | < .0001 |
|  | 11–20 | 37 | 50.7% | 18 | 51.4% | 55 | 50.9% |  |
|  | 21–30 | 29 | 39.7% | 2 | 5.7% | 31 | 28.7% |  |
|  | 31–40 | 4 | 5.5% | 1 | 2.9% | 5 | 4.6% |  |
|  | ≥41 | 3 | 4.1% | 0 | 0.0% | 3 | 2.8% |  |
| Difficulty of | Top 25% | 35 | 47.9% | 7 | 20.0% | 42 | 38.9% | 0.1233 |

|  |  |  |  |  |  |  |  |
| --- | --- | --- | --- | --- | --- | --- | --- |
| admission | Top 25%–50% | 20 | 27.4% | 8 | 22.9% | 28 | 25.9% |
|  | Bottom 25%–50% | 18 | 24.7% | 4 | 11.4% | 22 | 20.4% |
|  | Bottom 25% | 0 | 0.0% | 16 | 45.7% | 16 | 14.8% |
| Total |  | 73 | 100.0% | 35 | 100.0% | 108 | 100.0% |

### 3.2. Sex disparity among pharmacy professors in Japan and South Korea

Table 2 presents the population of Japan and Korea, the number of professors in pharmacy schools, and the number of professors per million population. A total of 1,965 pharmacy professors were identified in Japan and South Korea. Of these, 1,527 were from Japan and 438 were from South Korea, indicating that Japan had approximately 3.5 times as many professors in university pharmacy departments as South Korea. The estimated ratio of professors to million individuals in Japan was 12.28, compared with 8.51 in South Korea. After correction for population, this indicated that Japan had approximately 1.4 times as many professors as South Korea.

**Table 2.** Number of pharmacy professors per million individuals in Japan and South Korea.

| Classification | Japan | Korea |
| --- | --- | --- |
| Population* | 124,371,000 | 51, 474,900 |
| Number of professors | 1,527 | 438 |
| Number of professors per million individuals | 12.28 | 8.51 |

\*: United Nations. World Population Prospects 2024. https://population.un.org/wpp/data-sources

Regarding the sex distribution of professors, the proportion of female professors in Japan was 12.5% (191 of 1,527), while that in South Korea was 32.9% (144 of 438). This indicates that the proportion of female professors in both countries was low, falling below 50% (Table 3). A comparison of the number of female professors in the two countries revealed a significant disparity, with Japan exhibiting a markedly lower proportion of female professors than South Korea (*p* < .0001, chi-squared test).

**Table 3.** Sex distribution of pharmacy department professors in Japan and South Korea.

| Classification |  | Male |  | Female |  | Total |  | Chi-squared test |
| --- | --- | --- | --- | --- | --- | --- | --- | --- |
|  |  | Number of professors | % | Number of professors | % | Number of professors | % |  |
| Country | Japan | 1,336 | 87.5% | 191 | 12.5% | 1,527 | 77.7% | < .0001 |
|  | South Korea | 294 | 67.1% | 144 | 32.9% | 438 | 22.3% |  |
| Total |  | 1,630 | 83.0% | 335 | 17.0% | 1,965 | 100.0% |  |

### 3.3. Sex disparity by field of specialty and university background

A comparative analysis of the sex ratio of professors by specialty in Japan and South Korea revealed that the proportion of female professors in Japan was consistently lower than that in South Korea across all research fields, including basic and clinical research (Figure 1, Supplemental Table 1). In South Korea, 63.8% (44 of 69) of professors engaged in clinical research were women. Moreover, a comprehensive examination of clinical research in South Korea revealed that more than 50% of professors in the fields of pathology and drug therapy, as well as in the areas of law, systems, ethics, and clinical training, were women. Conversely, the proportion of female professors in Japan was less than 50% across all specialties, with the highest representation observed in the field of “laws, systems, and ethics,” which comprised 23.1% (9 of 39). Furthermore, the sex ratio of professors was examined by university background in both Japan and South Korea. The analyses, which considered factors such as the founder, faculty composition, location, and sex of students, revealed that the proportion of female professors was consistently lower in Japan than in South Korea. Regarding women’s universities in South Korea, approximately half of the professors were female (47.1%, 24 of 51). However, no categories in Japan had a proportion of female professors exceeding 50%.

**Fig 1.**
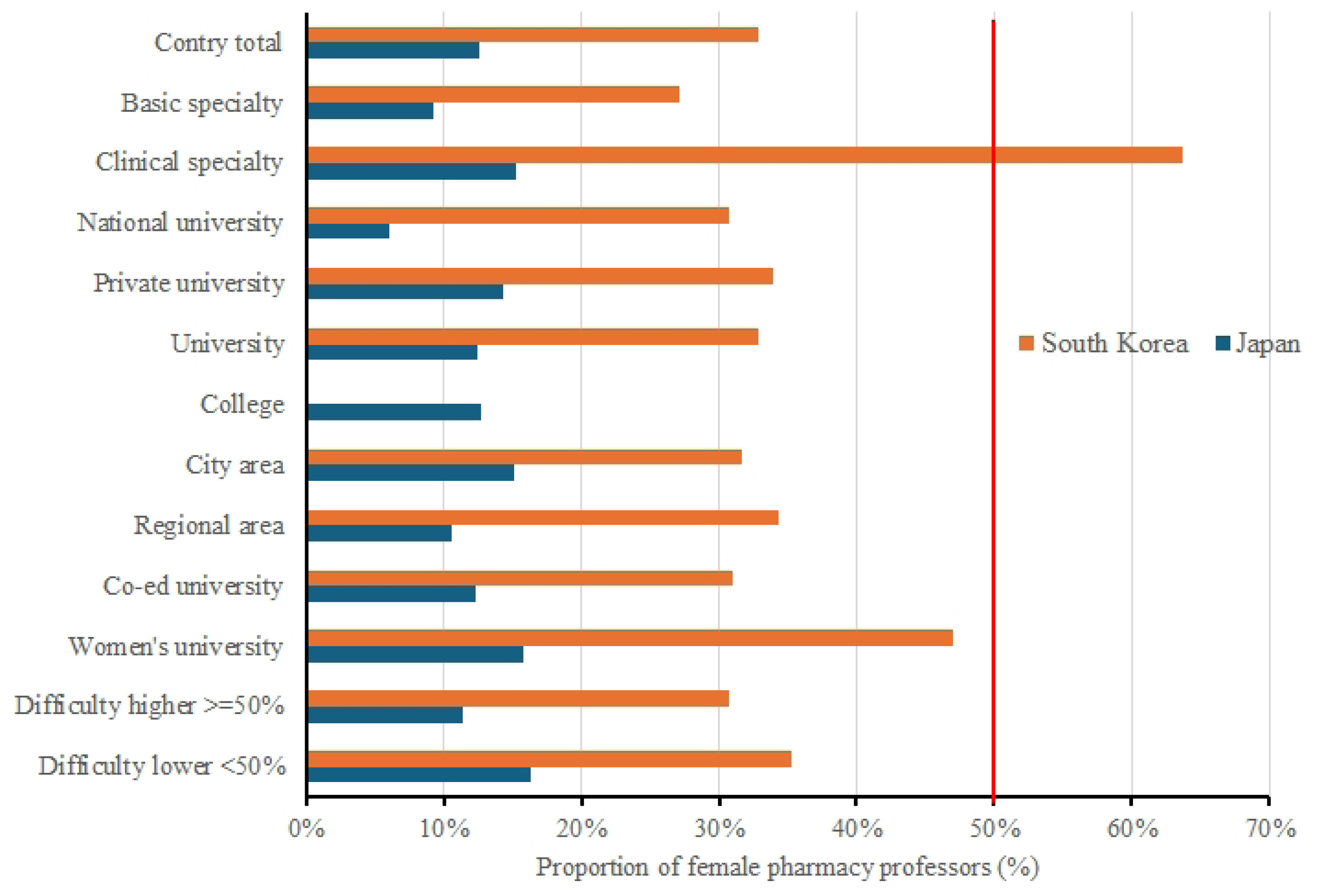
Proportion of female pharmacy professors, classified by stratification

### 3.4. Other factors

A major aim of university pharmacy departments is to contribute to success in the pharmacist national examination. In Japan, the pass rates of the national pharmacist examination by university were publicly available (https://www.mhlw.go.jp/content/11121000/001074357.pdf). Consequently, universities were classified according to their pass rates. Subsequently, an exploratory analysis was conducted to examine sex disparities in Japanese universities based on pass rates (Figure 2). Figure 2A illustrates the proportion of universities according to their national examination pass rate, while Figure 2B depicts the proportion of professors according to their national examination pass rate. The distribution of universities and professors by national examination pass rate revealed that the 90%–100% pass rate category comprised three universities (4.1%) and 66 professors (4.3%); the 80%–89% pass rate category comprised 27 universities (37.0%) and 592 professors (38.8%); the 60%–79% pass rate category comprised 22 universities (30.1%) and 444 professors (29.1%); the 40%–59% pass rate category comprised 17 universities (23.3%) and 357 professors (23.4%); and the ≤39% pass rate category comprised four universities (5.5%) and 68 professors (4.5%). No significant differences were observed between these categories regarding the proportion of universities and professors. Figure 2C illustrates the sex disparity among professors according to pass rate, with the proportion of female professors ranging between 8.6% in the 80%–89% pass rate category and 19.1% in the ≤39% pass rate category. Although the percentage of female professors was slightly higher in the lowest pass rate category, no clear trend was observed regarding sex disparity.

**Fig 2.**
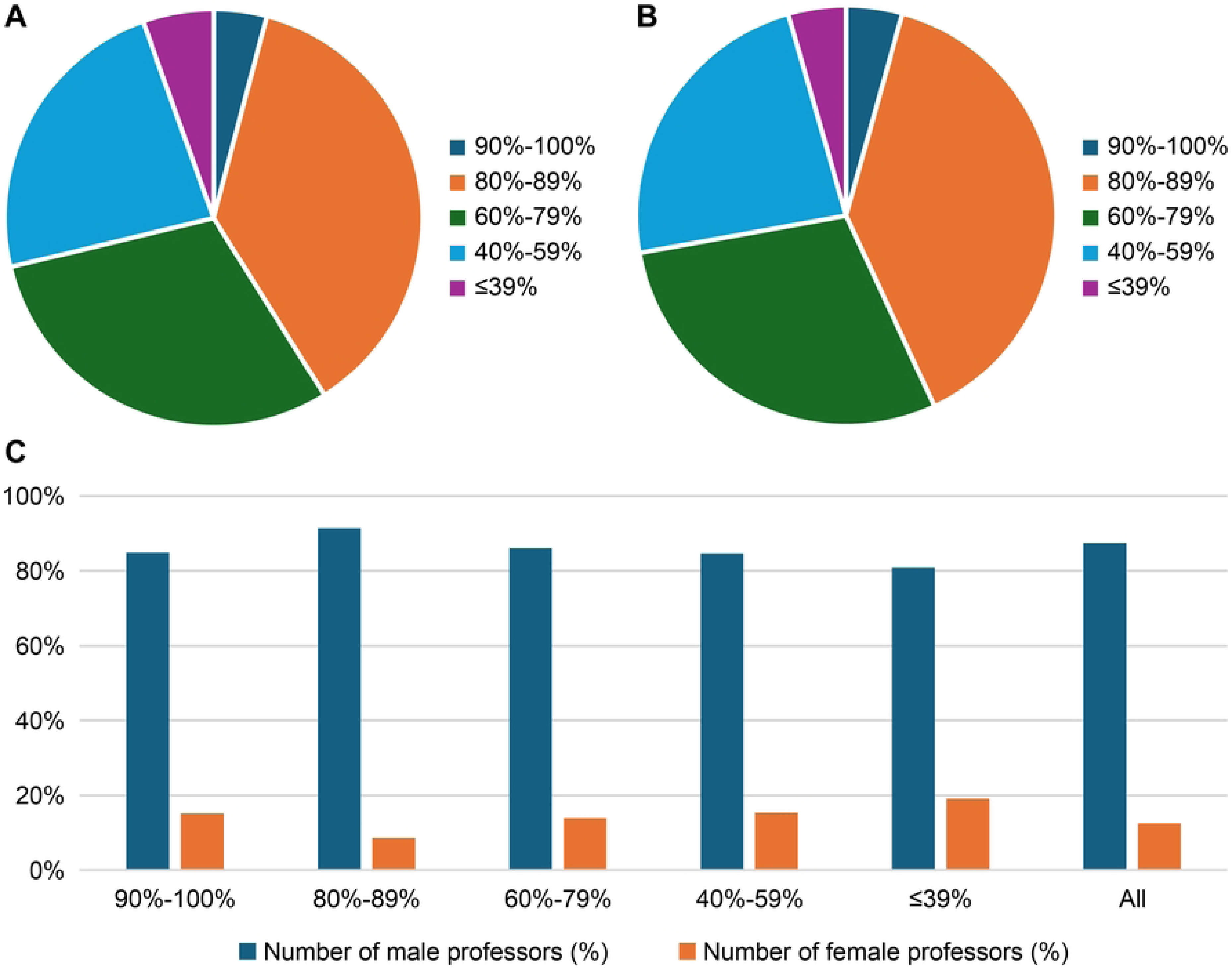
Sex disparity among professors in universities according to Japanese national examination pass rates

2A: Number of universities according to national examination pass rates (%); 2B: Number of professors according to national examination pass rates (%); 2C: Sex disparity among professors according to national examination pass rates (%)

## 4. Discussion

### 4.1. Previous studies on sex disparity among healthcare professionals and medical/pharmaceutical researchers

A paucity of previous studies examined sex disparity among university pharmacy professors. To date, no studies focused on sex disparity among pharmacy faculty in Japanese or South Korean universities. To the best of our knowledge, this study represents the inaugural investigation into this area. However, several previous studies examined sex disparities among medical researchers and healthcare professionals. For example, a study conducted in French hospitals on healthcare professionals revealed that a gender gap persists regarding access to directors in medicine and pharmacy but not in dentistry. Despite a trend towards gender equality over the past two decades, equality has not yet been achieved concerning access to the highest positions [14]. Furthermore, previous studies indicated a dearth of women in senior roles within the healthcare sector. This includes positions such as faculty members at medical centers [15], physician-scientists [16], hospital directors [17], and deans of medical schools [18]. Moreover, disparities have been documented concerning the number of healthcare professionals and their salaries [19]. Future studies on professors should include a meta-analysis of sex disparity among full professors as academic physicians 20]. Furthermore, disparities are not exclusive to the treatment of research subjects by researchers, educators, and healthcare professionals; women are also underrepresented among authors of high-impact papers [21], indicating a gender gap [22]. This gender gap has been reported in traditional medical research as well as in recent studies, such as studies on COVID-19 [23]. Additionally, previous findings suggest that women are subjected to bias in the conduct of research, as authors of papers, and in roles such as editor-in-chief [24] and the citation of papers [25]. This results in research themes pursued by women being deprioritized or excluded.

### 4.2. Previous studies on sex disparity in healthcare professions outside of the US and Europe

Sex disparity among researchers and healthcare professionals is not limited to developed countries such as Europe and the US. Reports of sex disparity have also originated from various other countries including Japan [24,26], South Korea [27], China [28], Peru [29], and Nigeria [30]. This suggests that sex disparity among medical researchers and healthcare professionals is a global issue.

### 4.3. Gender Gap Index

This study examined sex disparities among professors in university pharmacy departments in Japan and South Korea. The results indicated that the proportion of female professors was less than 50% in both countries; however, Japan had a significantly lower number of female professors than South Korea. According to the 2024 World Economic Forum Gender Gap Index [1], Japan was ranked 118th and South Korea was ranked 100th. The findings of this study, which indicated a greater sex disparity in Japan than in South Korea, are consistent with the gender gap index.

### 4.4. Sex disparity among university pharmacy department professors in Japan and South Korea

Regarding the specialties of professors, female professors were more likely to specialize in clinical research than in basic research. This trend was particularly pronounced in South Korea; however, both countries had a higher proportion of female professors in the areas of “Law, Systems, and Ethics,” “Disease and Drug Therapy,” and “Clinical Training.” An explanation for this may be the underlying perception that women are less adept at science subjects and that men excel in these areas from an early age. Addressing such preconceptions and stereotypes and improving education from early childhood is crucial for reducing sex disparity. Additionally, women who become professors may have gained clinical experience in hospitals or pharmacies before returning to academia. In such cases, women may take longer breaks from work than men owing to marriage and childbirth, which may hinder their career progression [2]. This may explain the lower number of women in the highest academic positions, such as professors. Previous statistics indicated that there are more female students and practitioners in pharmacy than their male counterparts [9,10], Therefore, increasing the number of female professors in pharmacy departments and promoting diversity is critically important.

## 5. Limitations

This study had some limitations. First, the sex determination was predominantly based on first names and/or photographs, which may have resulted in erroneous classifications. Second, the study was limited in its scope, focusing exclusively on professors and not encompassing all academic disciplines. This may have resulted in an incomplete representation of the actual situation pertaining to the entire academic staff. In particular, some newly established pharmacy departments in South Korea may not yet have any professors. Third, the classification of professors’ specialties was based on the criteria used for Japan’s national exams, which may not align with South Korea’s standards. Fourth, the difficulty of university admission in South Korea was classified based on personal judgment rather than official national data, which may differ from the actual level of difficulty.

## 6. Conclusions

This study examined sex disparity among 1,965 professors from 73 pharmacy schools in Japan and 35 university pharmacy departments in South Korea. Of the total number of professors surveyed, 1,527 were from Japan and 438 from South Korea. Japan had approximately 1.4 times as many professors as South Korea per million population. In Japan, the proportion of female professors was 12.5% (191 of 1,527), whereas the proportion in South Korea was 32.9% (144 of 438). Both countries exhibited a female professor population of less than 50%. Both Japan and South Korea demonstrated sex disparity among university pharmacy department professors. However, Japan exhibited a significantly lower proportion of female professors than South Korea. This indicates that the advancement of women in Japanese pharmacy faculties is lagging behind that in South Korea. The greater disparity in Japan is consistent with the findings of the World Economic Forum’s gender gap index. Furthermore, the higher proportion of female professors specializing in clinical research in South Korea may indicate that women with clinical experience in hospitals and pharmacies are more likely to become professors in university pharmacy departments. This study’s findings identified the current state of sex disparities among faculty members at pharmacy schools in Japan and Korea and emphasized future challenges for promoting diversity and equality in academia and the pharmacy profession.

## Data Availability

Data available on request from the researchers after approval by the author. Basically, the data could be available, however the data in this study was created originally by the authors, and the authors would like to know who will use the data and how.

## ACKNOWLEDGMENTS

We would like to thank Editage (www.editage.com) for English language editing.

## Ethical Considerations

This study did not require institutional review board approval or patient informed consent because it was based on publicly available information and included no patient records.

## Data availability

Data available on request from the researchers after approval by the author.

## References

1. World Economic Forum. Global gender gap 2024, insight report; 2024. [Cited January 12, 2025]. Available from: https://www3.weforum.org/docs/WEF_GGGR_2024.pdf.

2. The Japan times. Japan makes gains in political empowerment in gender equality report. [Cited January 12, 2025]. Available from: https://www.japantimes.co.jp/news/2024/06/12/japan/society/japan-gender-gap/.

3. The Korea times. Korea’s gender wage gap worst among 33 OECD countries: report. [Cited January 12, 2025]. Available from: https://www.koreatimes.co.kr/www/biz/2024/08/602_370268.html.

4. Yousaf R, Schmiede R. Barriers to women’s representation in academic excellence and positions of power. Asian J Ger Eur Stud. 2017;2. doi: 10.1186/s40856-017-0013-6.

5. Hori RS. Progress and problems of gender equality in Japanese academics and geosciences. Adv Geosci. 2020;53: 195–203. doi: 10.5194/adgeo-53-195-2020.

6. Homma MK, Motohashi R, Ohtsubo H. Japan’s lagging gender equality. Science. 2013;340: 428–430. doi: 10.1126/science.340.6131.428-b.

7. Gender equalty bureau cabinet office in Japan. Expansion of women’s participation in policy and decision-making processes in all fields in society. [Cited January 12, 2025]. Available from: https://www.gender.go.jp/english_contents/mge/process/index.html.

8. Ministry of Gender Equality and Family. Development and implementation of the “The 2nd Framework Plan for Gender Equality Policies”. [Cited January 12, 2025]. Available from: https://www.mogef.go.kr/eng/pc/eng_pc_f001.do.

9. Devraj R, Warholak T, Planas LG. Seeking gender equity in pharmacy academia. Am J Pharm Educ. 2023;87: ajpe9050. doi: 10.5688/ajpe9050.

10. Bissell BD, Johnston JP, Smith RR, Newsome AS, Thompson Bastin ML, Abdul-Mutakabbir J, et al. Gender inequity and sexual harassment in the pharmacy profession: Evidence and call to action. Am J Health Syst Pharm. 2021;78: 2059–2076. doi: 10.1093/ajhp/zxab275.

11. Maeda H, Hyunseo S, Hashimoto M, Han E, Ohno E. Gender differences in pharmacy professors in Japan and Korea. Lancet Reg Health West Pac. 2025; *in press*.

12. Vandenbroucke JP, von Elm E, Altman DG, Gøtzsche PC, Mulrow CD, Pocock SJ, et al. Strengthening the reporting of observational studies in epidemiology (STROBE): Explanation and elaboration. Epidemiology. 2007;18: 805–835. doi: 10.1097/EDE.0b013e3181577511.

13. Ministry of Health and Welfare. Japan. National Pharmacist Examination Page. Available from: https://www.mhlw.go.jp/stf/seisakunitsuite/bunya/kenkou_iryou/iyakuhin/yakuzaishi-kokkashiken/index.html (in Japanese) [Cited January 12, 2025].

14. Le Boedec A, Anthony N, Vigneau C, Hue B, Laine F, Laviolle B, et al. Gender inequality among medical, pharmaceutical and dental practitioners in French hospitals: Where have we been and where are we now? PLOS One. 2021;16: e0254311. doi: 10.1371/journal.pone.0254311.

15. Franks AM, Calamur N, Dobrian A, Danielsen M, Neumann SA, Cowan E, et al. Rank and tenure amongst faculty at academic medical centers: A study of more than 50 years of gender disparities. Acad Med. 2022;97: 1038–1048. doi: 10.1097/ACM.0000000000004706.

16. Ward HB, Levin FR, Greenfield SF. Disparities in gender and race among physician-scientists: A call to action and strategic recommendations. Acad Med. 2022;97: 487–491. doi: 10.1097/ACM.0000000000004224.

17. Watari T, Gupta A, Hayashi M, Mizuno K, Nakano Y, Tokuda Y, et al. Characteristics of Medical School deans and University Hospital Directors in Japan. JAMA Netw Open. 2024;7: e2351526. doi: 10.1001/jamanetworkopen.2023.51526.

18. Gottlieb AS, Roy B, Herrin J, Holaday LW, Weiss J, Salazar MC, et al. Why are there so few women Medical School deans? Debunking the myth that shorter tenures drive disparities. Acad Med. 2024;99: 63–69. doi: 10.1097/ACM.0000000000005315.

19. Steffler M, Chami N, Hill S, Beck G, Cooper SC, Dinniwell R, et al. Disparities in physician compensation by gender in Ontario, Canada. JAMA Netw Open. 2021;4: e2126107. doi: 10.1001/jamanetworkopen.2021.26107.

20. Marhoffer EA, Ein-Alshaeba S, Grimshaw AA, Holleck JL, Rudikoff B, Bastian LA, et al. Gender disparity in Full Professor Rank among academic physicians: A systematic review and meta-analysis. Acad Med. 2024;99: 801–809. doi: 10.1097/ACM.0000000000005695.

21. Fathy CA, Cherkas E, Shields CN, Syed ZA, Haller JA, Zhang QE, et al. Female editorial authorship trends in high-impact ophthalmology journals. JAMA Ophthalmol. 2021;139: 1071– 1078. doi: 10.1001/jamaophthalmol.2021.3027.

22. Yalamanchali A, Zhang ES, Jagsi R. Trends in female authorship in major journals of 3 oncology disciplines, 2002-2018. JAMA Netw Open. 2021;4: e212252. doi: 10.1001/jamanetworkopen.2021.2252.

23. Misra V, Safi F, Brewerton KA, Wu W, Mason R, Chan AW, et al. Gender disparity between authors in leading medical journals during the COVID-19 pandemic: A cross-sectional review. BMJ Open. 2021;11: e051224. doi: 10.1136/bmjopen-2021-051224.

24. Harada K, Ozaki A, Murayama A, Tsuji A, Miyachi T, Yamamoto K, et al. Woman Editors-in-Chief of English-Language Medical Journals Published by the Japanese Professional Medical Associations. JMA J. 2022;5: 114–117. doi: 10.31662/jmaj.2021-0008.

25. Shamsi A, Lund B, Mansourzadeh MJ. Gender disparities among highly cited researchers in biomedicine, 2014-2020. JAMA Netw Open. 2022;5: e2142513. doi: 10.1001/jamanetworkopen.2021.42513.

26. Akazawa S, Fujimoto Y, Sawada M, Kanda T, Nakahashi T. Women physicians in academic medicine of Japan. JMA J. 2022;5: 289–297. doi: 10.31662/jmaj.2021-0116.

27. Lee MJ, Kim C. Breaking the gender gap: A two-part observational study of the gender disparity among Korean academic emergency physicians. J Prev Med Public Health. 2020;53: 362–370. doi: 10.3961/jpmph.20.286.

28. Zheng X, Vastrad S, He J, Ni C. Contextualizing gender disparities in online teaching evaluations for professors. PLOS One. 2023;18: e0282704. doi: 10.1371/journal.pone.0282704.

29. Amaya E, Mougenot B, Herrera-Añazco P. Gender disparities in scientific production: A nationwide assessment among physicians in Peru. PLOS One. 2019;14: e0224629. doi: 10.1371/journal.pone.0224629.

30. Folayan MO, Olowokeere A, Lusher J, Aina O, Gascon A, Martínez-Pérez GZ. A qualitative insight into researchers’ perceptions of gender inequality in medical and dental research institutions in Nigeria. PLOS One. 2023;18: e0283756. doi: 10.1371/journal.pone.0283756.

